# An Open Access Collaborative Global Virtual Biorepository System (VBS): A Delphi Consensus

**DOI:** 10.1101/2025.10.20.25338417

**Authors:** Amy Price, Layla Abdulbaki, Julia Poje, Judith Giri, Geoffrey Winstanley, Zoe Steinberg, Thomas Jaenisch, May Chu

**Author notes:** Corresponding Author, Professor May Chu word count 3,180 (minus abstract and references).

## Abstract

**Background:** The re-emergence of Zika virus, the novel coronavirus, and new influenza strains highlighted the urgent need for an organized forum to access specimens for advancing research, diagnostics and vaccine development. A Virtual Biorepository System (VBS) was proposed to connect communities and samples, addressing specimen-sharing challenges while promoting inclusion and equity.

**Aim and Objectives:** To enable equitable contribution, sharing, and access to specimens while addressing the complexities involved. The objective is to build stakeholder consensus on prioritized actions, tools, and responsibilities to establish a functional and practical VBS.

**Methods:** A Delphi process was conducted online to gather stakeholder input and develop a prioritized action plan for the VBS. Respondents were recruited using snowball sampling, and questions were informed by participatory workshops, feedback, outreach efforts, and discussions to identify gaps and best practices.

The Delphi process consisted of two rounds: an initial ranking informed by stakeholder inputs and a second round of refining priorities based on panel feedback. Consensus threshold was defined as ≥70% of responses rating an item ≥4 and excluded if an item had ≤15% response or was rated ≤2 on a 1–5 Likert scale. Advisory board discussions supplemented the virtual process to capture nuances, while qualitative data were analyzed using participatory action research methods.

**Findings:** The Delphi process identified critical gaps and priorities in biorepository systems. Consensus was reached for 22 out of 30 items. Items 20 of 26 in Round-1, with 2 of 4 additional items in Round-2 where consensus was reached on 2 items addressing benefit-sharing challenges (73%) and prioritizing whole blood, plasma, and serum samples (95%).

Respondents overwhelmingly supported a hybrid VBS model combining federated and centralized systems (83%) cost recovery for services (100%) Some respondents suggested an equitable fee structure using a sliding scale to subsidize capacity building in low-and middle-income countries (95%).

**Conclusions:** Respondents strongly supported a VBS to enable equitable access to specimens, services, and benefits. They emphasized the need for clarity on benefit-sharing, regulatory, and ethical frameworks to ensure effective implementation and global inclusivity. A fair and equivalent system to enhance participation and operational continuity is being discussed.

## Introduction

Access to high-quality, well-characterized specimens that reflect global diversity, including populations from low- and middle-income countries (LMICs), is critical for advancing research, diagnostics, and vaccine development [1]. Despite the evident need during outbreaks such as Zika (2016) and Coronavirus Infectious Disease-2019 (COVID-19), and concerns around evolving pathogen threats, a sustainable and trusted global resource for these samples has yet to be realized. While the concept is not new, progress has been hindered by challenges in regulatory and ethical implementation, as well as concerns over tangible benefits and long-term sustainability, which have discouraged partners from advancing these efforts [1]. Disparities in diagnostic kit performance across high- and low-income settings underscore the importance of globally representative specimens [2,3]. The COVID-19 pandemic, in particular, exposed delays in accessing essential samples, and hampered timely research and development [4].

Historically, international collections for infectious diseases have been centralized within large, well-funded institutions and biorepositories, often excluding local contributors and geographically diverse populations [5]. The pandemic amplified inequities in diversity, equity, and inclusion within these systems [2,3]. To address these challenges, we propose a virtual biorespository system (VBS) that connects local repository hubs for efficient, equitable, and accessible sample sharing [1].

The VBS is designed to be agile and inclusive, combining the strengths of grassroots initiatives with sensitivity to local contexts as *an additional repository type* to the standard centralized systems. By reducing the need for a centralized multinational facility, the VBS reduces overhead but then could be limited by the type and use of its specimens, so we needed to find concensus on how to advance equity while defining the types of accessible samples with simple storage needs that a global partner with limited resources could join the effort.

This study employs an international online Delphi to achieve consensus and solve challenges collaboratively [6]. The Delphi method offers several advantages:

1. Empowers under-resourced populations by fostering inclusion and reimagining global biorepository systems.
2. Amplifies local voices to define the benefits and sustainability of the VBS.
3. Creates a roadmap for addressing end-user priorities.
4. Reduces power imbalances through anonymous contributions.
5. Accommodates cultural and geographic diversity across respondents.

The need for a systematic, inclusive approach to specimen access is urgent, particularly as global health threats like Zika, emerging coronaviruses, and influenza variants resurface [5,7]. Biorepositories are essential for providing qualified, annotated samples that advance diagnostics, disease prediction, and prevention efforts [1,4].

Our goal is to develop actionable recommendations for building and maintaining a virtual biorepository while fostering ongoing consensus as the initiative evolves. This grassroots effort aims to facilitate scientific exchange, equitable specimen sharing, and global collaboration as a public good. By aligning with successful international partnerships, the VBS seeks to be an inclusive, additive, and sustainable model for a multinational community. Using the Delphi method we plan to identify practices that promote biorepository availability, good governance, and long-term sustainability [8]. Our protocol and supplementary materials are available on the Open Science Framework and in the supplementary materials [9].

## Methods

### Preparation Work

The research team collected information from listserv contacts, conducted surveys and interviews [1], held workshops, searched the literature, set up websites, and connected with others in the field, including biorepository experts, public health practitioners, policymakers, industry leaders, and funders, students and members of the public. The research team conducted internal audits to systematically evaluate the effectiveness and alignment of our activities with the project’s goals. These audits reviewed the outcomes of workshops, and stakeholder engagements, assessed gaps in literature and expert input, and identified actionable next steps [1]. Through this reflective process, we deliberated and concluded that an online Delphi could offer a practical approach to bridge cultural, linguistic, and geographical divides, foster autonomous local consensus [10], and ensure cost-effectiveness [11]. Figure 1 shows the timeline for the VBS and the steps we used to prepare for the Delphi.

**Figure 1.**
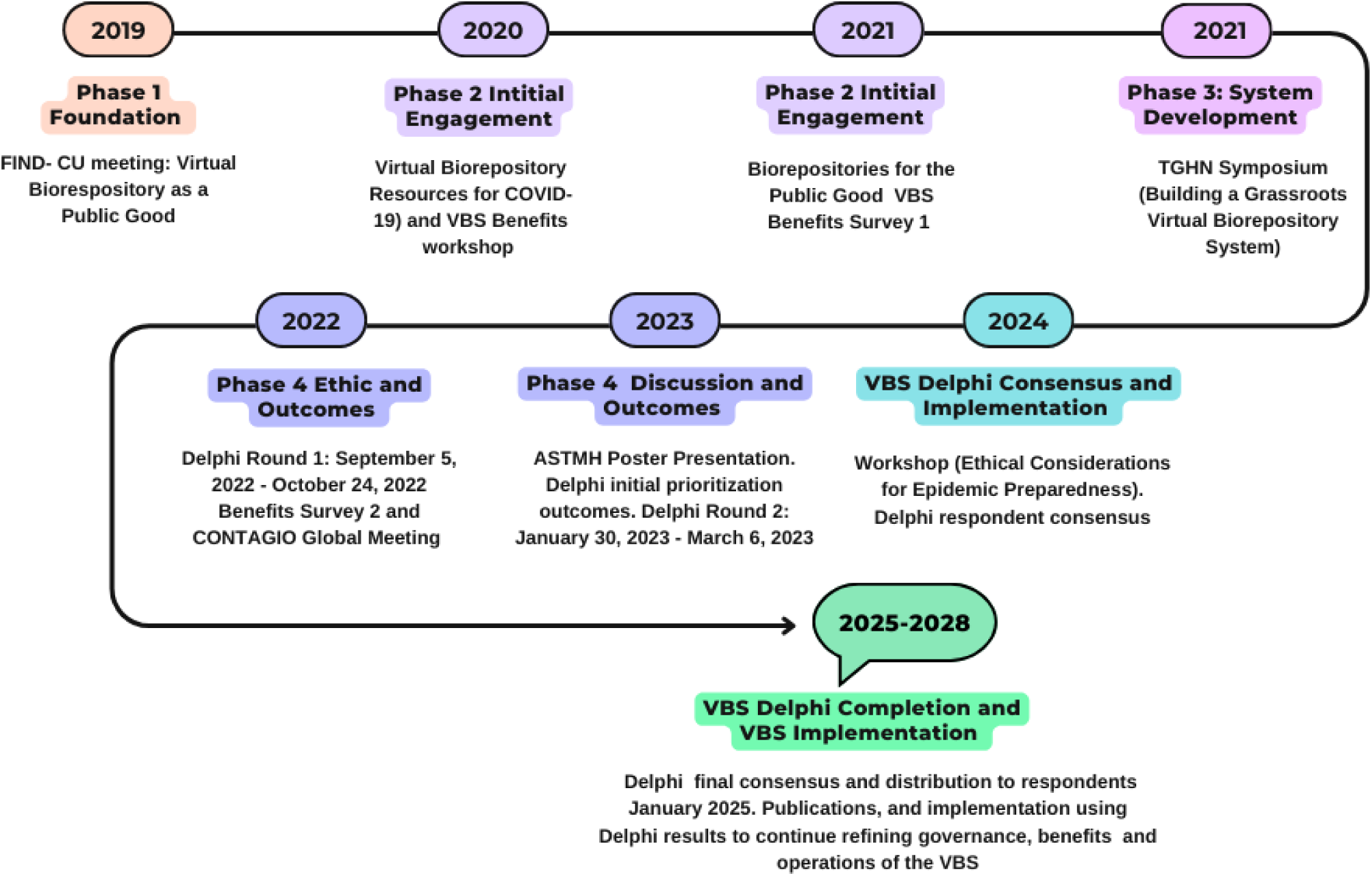
Preparatory work and Delphi Timeline. FIND, Foundation for Innovative Diagnostics; CU, University of Colorado; VBS, Virtual Biorepository System; TGHN, The Global Health Network; CONTAGIO, Cohort Network to be Activated Globally in Outbreaks; ASTMH, American Society for Tropical Medicine and Hygiene.

### The Delphi Process

The Delphi is a structured, iterative communication tool designed to facilitate consensus-building, forecasting, or shared decision-making. Through a series of rounds, whether delivered onsite or by using virtual survey software, respondents evaluate and prioritize presented questions, typically participating in at least two rounds [8]. The Delphi was offered online through a secure interface. After each round, participants received an anonymous summary of the group’s inputs, enabling them to refine and prioritize their previous responses based on others’ feedback. This iterative process narrows the range of variation until common ground is reached. The underlying principle is that structured group input yields more accurate and actionable results than unstructured discussions [6].

### Aims and Objectives

We aimed to identify prioritized actions, tools, and responsibilities necessary to establish a functional VBS. Specifically, our objectives were to:

1. Develop actionable, consensus-based recommendations for building and maintaining an inclusive, diverse, and equity-driven VBS.
2. Generate ongoing findings as the project evolves to ensure continued alignment with stakeholder needs and global priorities.

### Study Design

The VBS initiative was informed by global webinars, workshops, and feedback sessions involving participants from Africa, Latin America, Southeast Asia, and Europe. Contributors included biobank experts, blood bank consultants, public health specialists, researchers, and diagnostics manufacturers.

To shape grassroots, equitable approaches for specimen sharing, we employed a mixed-methods, sequential participatory action research design [12] centered around a multiple-round online Delphi. This process was built on insights from workshops, ongoing sample collection efforts, and stakeholder discussions to frame the Delphi questions. We piloted the questions based on key needs identified from feedback from stakeholders and consultative workshops [1], which highlighted three target areas:

1. **Types of specimens to share**: Identifying priority specimens for global research and diagnostics.
2. **Solutions to barriers**: Addressing logistical, regulatory, and ethical practices and cultural challenges in specimen sharing.
3. **Benefits and sustainability**: Articulating the value and long-term viability of equitable specimen-sharing practices.

The Delphi process engaged all stakeholders to rank, refine, and reach a consensus on these critical issues. Figure 2 provides a visual representation of the Delphi methodology and process.

**Figure 2.**
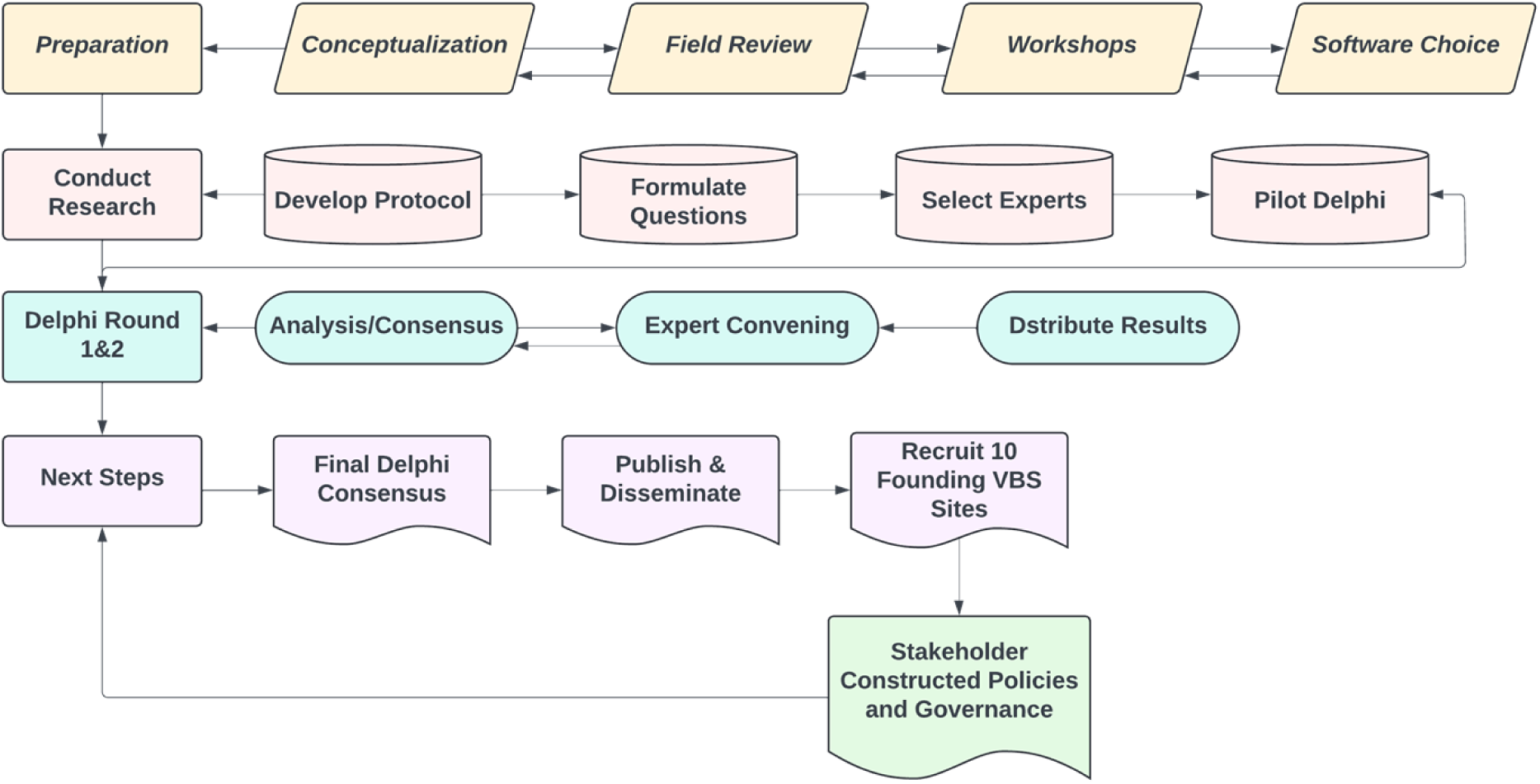
Virtual Biorepository Online Delphi Process

### Advisory Group and Steering Group

The Delphi development was guided by an expert advisory group specializing in infectious diseases, diagnostics manufacturing, and research methodology. The advisory group provided strategic oversight for the study design and its implementation. The advisory group and the researchers, together, formed the steering group to make decisions on the process. To ensure clarity, the Delphi questions were also reviewed by graduate students and advocates.

### Recruitment

Fifty-seven respondents were recruited using snowball sampling to ensure a diverse and inclusive panel. Stakeholders included public health experts, researchers, clinicians, community members, policymakers, regulators, and industry representatives, as well as respondents from six continents.

### Online Delphi Rounds

1. **Round-1 (Exploratory Phase)**: Respondents scored and commented on 26 initial items and were invited to propose additional questions. Items added by respondents were reviewed and coded by two research team members to ensure relevance.
2. **Round-2 (Evaluative Phase)**: Respondents reviewed results, including advisory group comments from Round-1, and voted again on unresolved items. Respondents scored the 4 additional items proposed during Round-1.

A consensus was reached after two rounds, and the results were reviewed and accepted by the advisory group. All results, including respondent comments with identifiers removed were compiled and shared with participants. Consensus threshold was defined as ≥70% of responses rated an item ≥4 and excluded if an item had ≤15% response or was rated ≤2 on a 1–5 scale.

### Delphi Response Preparation

The responses were scored and prioritized using a Likert scale, which for our questionnaire, varied between 1-5 points [13]. Respondents rated their level of agreement with each statement, with higher scores reflecting stronger agreement. This approach enabled a nuanced understanding of consensus among experts participating in the Delphi process. To further enrich the findings, qualitative content analysis was conducted. Five researchers (AP, LA, MC, JG, and JP) independently reviewed and categorized the textual responses provided by respondants, identifying themes, key concepts, and recurring opinions [6,11]. They reached a consensus on these interpretations, and the de-identified comments and the analysis were presented to the steering group and then the respondents. This contextual guidance informed the development of round two [14].

This method proved valuable in assessing how consensus evolves by considering levels of agreement or disagreement expressed in the comments alongside quantitative data, such as mean ratings or consensus thresholds. The analysis and de-identified feedback were shared with respondents after the first and second rounds, fostering transparency and enabling a reflective and shared understanding of the group’s perspectives. All respondents and the steering group were presented with Round-2 results. Following the Delphi rounds, a consensus conference was held via video conferencing with the advisory group to finalize recommendations and define key next steps.

### Ethical Considerations

Ethical approval was obtained from the University of Oxford Central University Research Ethics Committee (CUREC, #R58550/RE001) [15]. The Delphi was launched September 5, 2022, and results were completed and distributed to our participants in January 2025. (Please see Figure 1). The need for written or proxy signed consent was waived by the ethics committee. Clicking “Agree” to the consent conditions after reviewing the consent questions was considered informed consent. A copy of the questions is included in the supplementary materials. Contributors to the research, at their discretion, could be named as co-authors in the publication while maintaining the confidentiality of their data. Authors disclosed competing interests, adhering to ICMJE guidelines [16]. All data was securely stored online with restricted access limited to project-dedicated personnel. This study was determined not to constitute Human Subjects Research and was classified as minimal risk, quality improvement, and therefore not subject to full IRB review. Participants were informed of their right to withdraw at any time without penalty and were reassured that identifying data would be removed. Participants consented to the use of data that had already been collected and aggregated, with the understanding that such data would not be withdrawn. Delphi panel members who also contributed to the publication were, at their discretion, included as named contributors. However, all personal data were de-identified for analysis. This information was included for participants in the introductory section of the Delphi study and formed part of the participant agreement. The study underwent independent scientific review. Only Collaborative Institutional Training Institute (CITI)-trained research staff had access to identifiable data. Authors did not have access to identifiable data. The option to share an email address was added for internal demographics and to allow participants to (a) receive a copy of the publication and/or (b) be acknowledged by name if they wished. All participants were assigned a number that was accessible only to designated CITI-certified research staff. No personal data were, or will be, shared with third parties. All data are stored on secure, university-managed servers protected by firewalls.

### Data Collection and Research Reporting

Invitation letters, recruiting materials, forms, and Delphi questions were piloted by volunteers internally and externally. The Checklist for Reporting Results of Internet E-Surveys (CHERRIES) [17] guideline was used to structure and report the survey aspects of the study. The ACcurate COnsensus Reporting Document (ACCORD) was used to guide our methods for reporting the Delphi process [14].

### Patient and Public Involvement

The respondents and authorship team spanned multiple continents and brought diverse perspectives to the study. This included partners and stakeholders with an interest in the VBS, none of the respondents constituted patients. Additionally, the study engaged research, public health, and medical students, as well as members of the public, who actively contributed to the study design, protocol development, Delphi question testing, data analysis, and manuscript co-authorship.

## Results

### Quantitative Analysis

The Delphi study identified gaps and priorities in biorepository systems. Consensus was reached for 22 of 30 items: 20 of 26 in Round-1, and 2 of 4 additional items in Round-2 (see Tables 1–2). Both quantitative and qualitative data on stakeholder priorities for the VBS were collected. In Round-2, the advisory group and researchers elected to clarify the wording of items related to benefits and presented these questions again; however, this area still did not reach consensus. Items without consensus were excluded from further consideration. To present results, we used descriptive statistics and conducted a qualitative analysis of participant comments across both rounds. Respondents were provided with quantitative results and all unedited comments from the previous round. The items listed below represent those that reached consensus by percentage. To assess whether respondents would change their replies as they go through the questions, we repeated the question: Rank support for combined federated and centralized systems (see Question A, 37/47, 78%) and received 34/41 (83%) equivalent value replies upon requestioning. *(Complete results, including de-identified comments, response distributions, and items that did not reach consensus, are available in the supplementary materials)*.

**Table 1.**
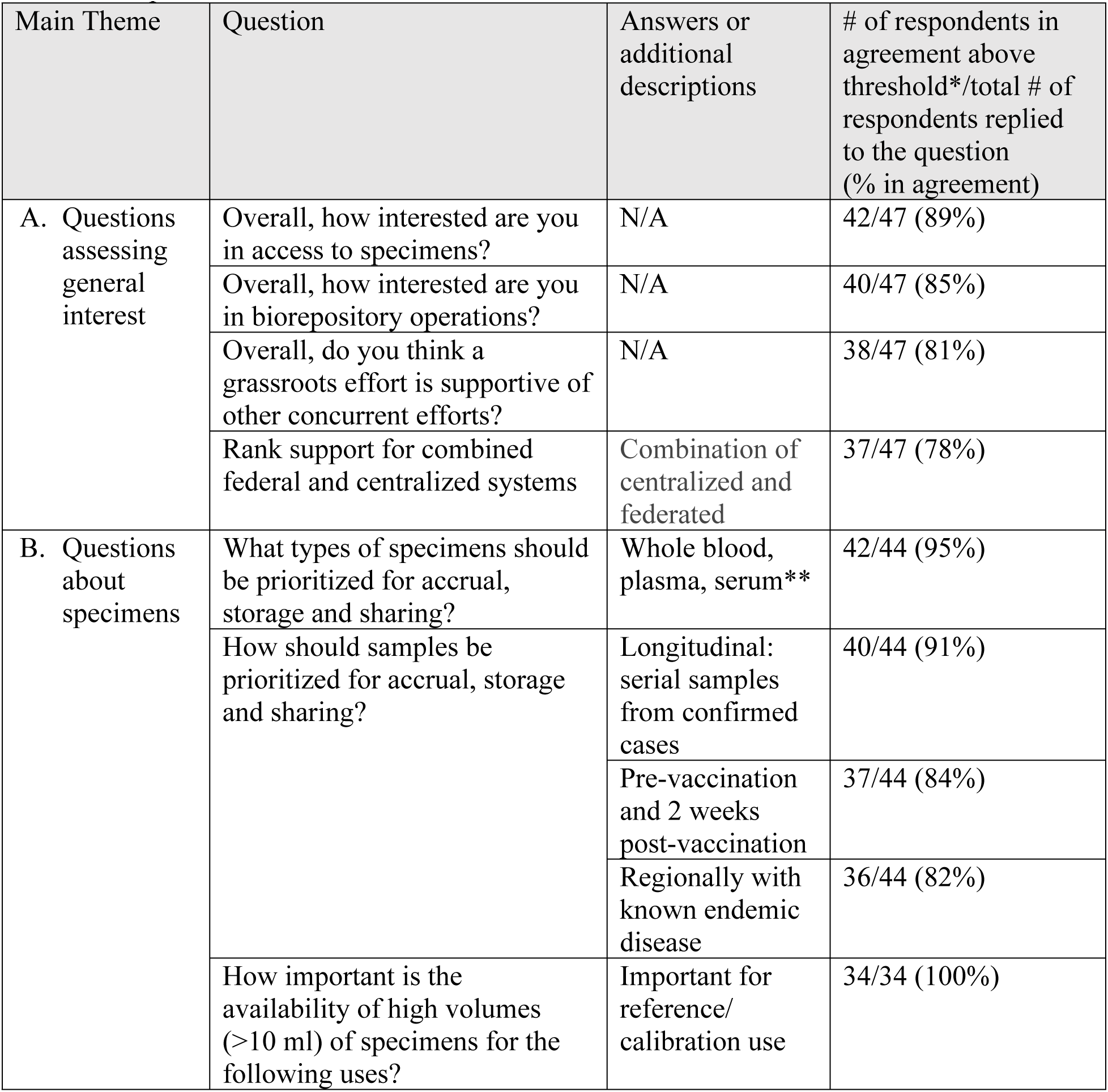
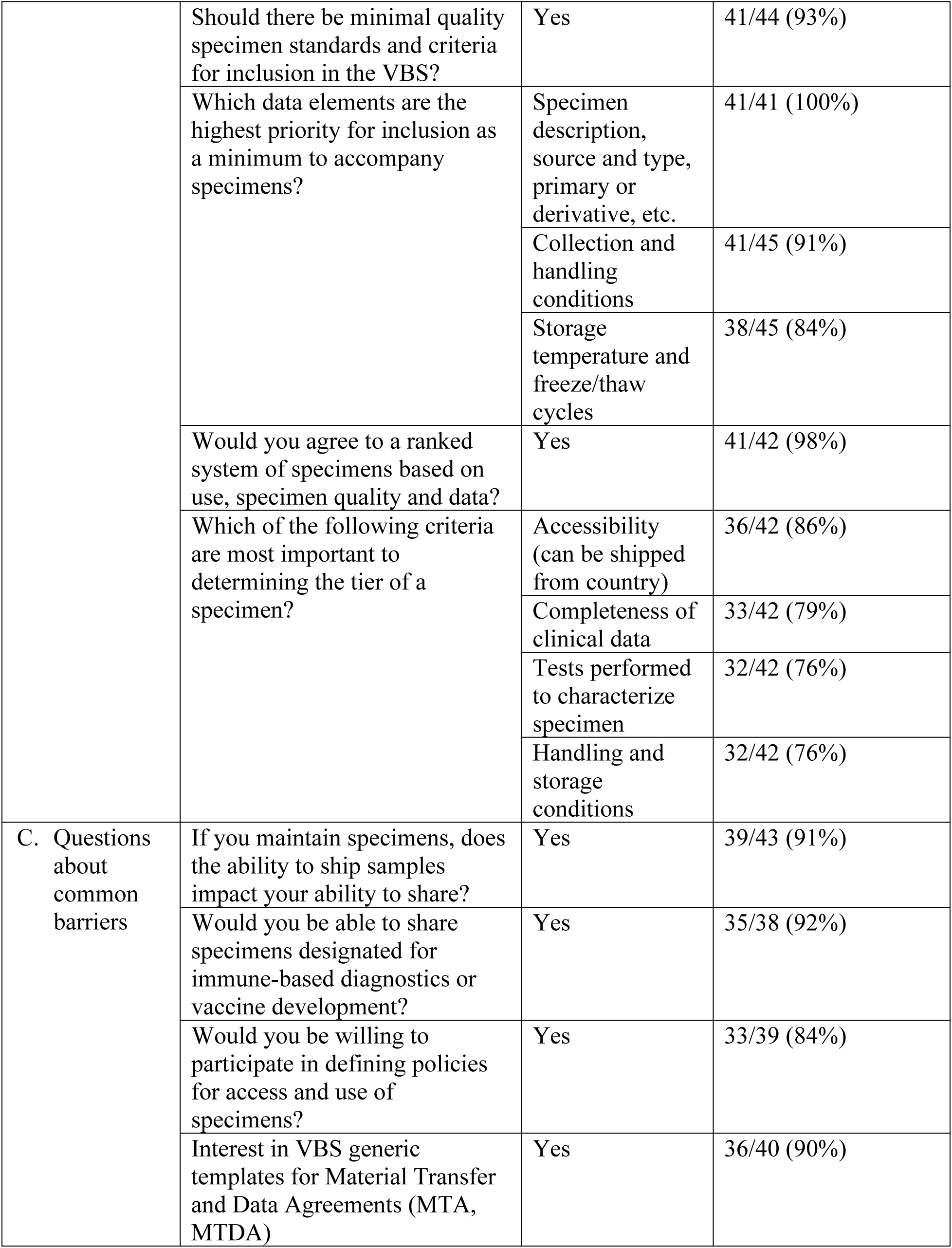
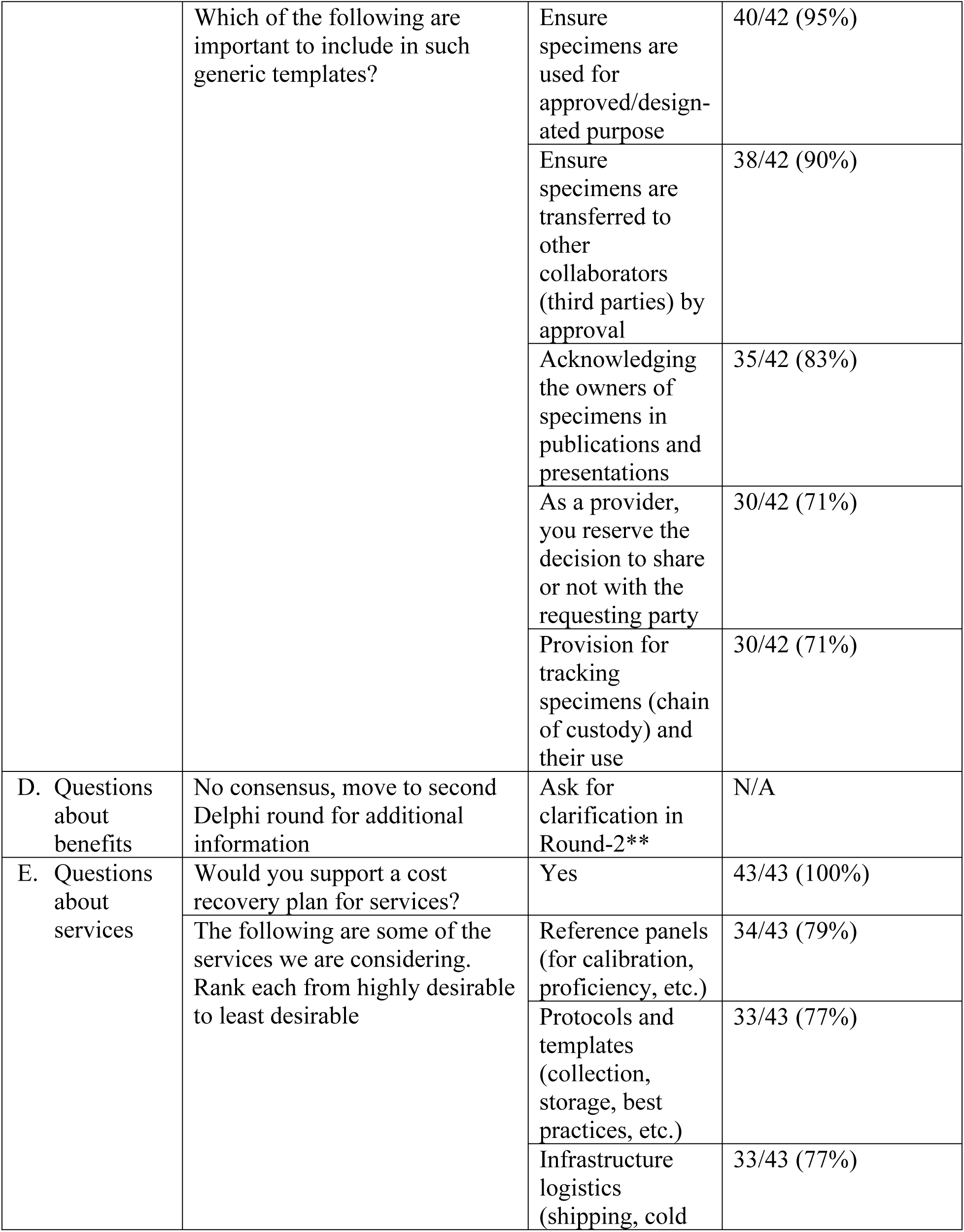
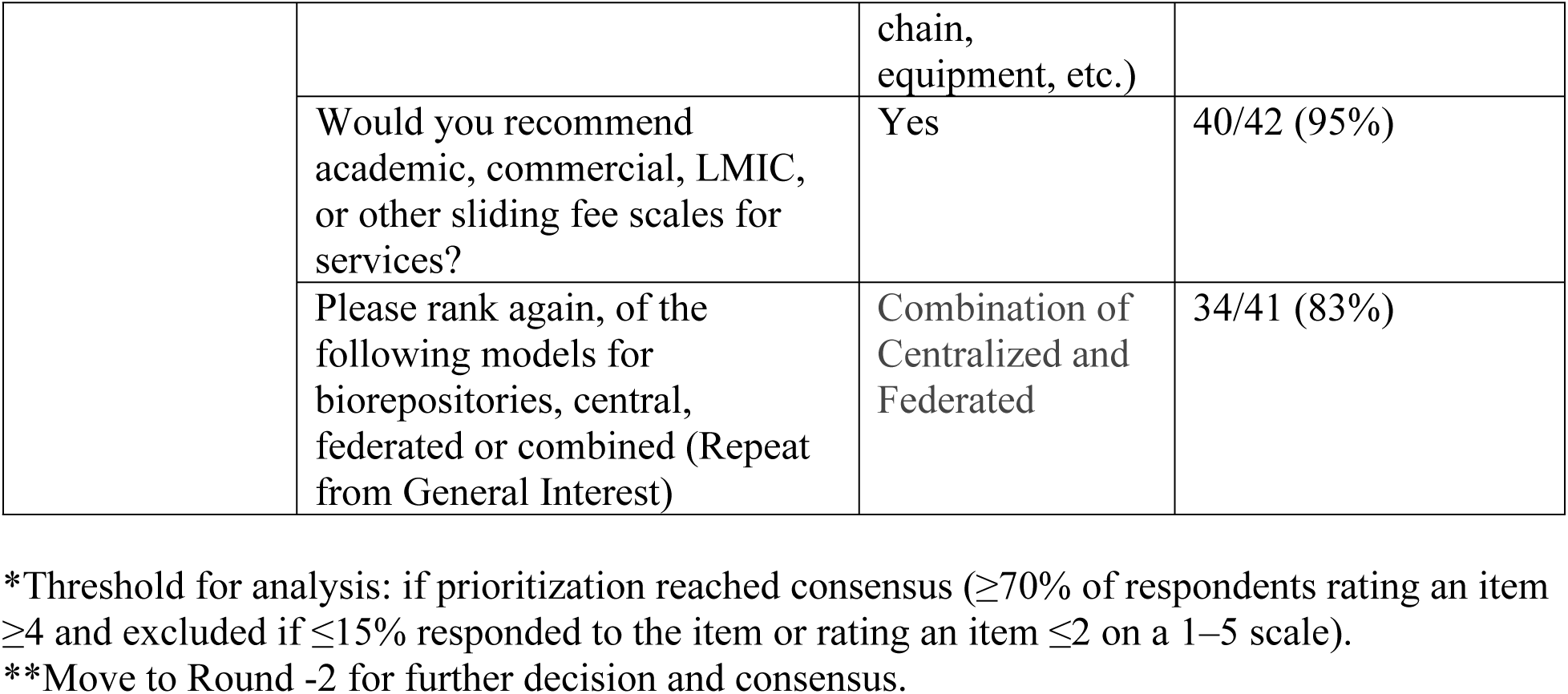
Delphi Results Round-1

**Table 2.** VBS Consensus Round-2

| Main Theme (from Table 1) | The 2 questions that did not receive consensus in the first round now posed to the second round for clarification | Answers to the questions or additional descriptions | # of respondents in agreement above threshold*/total # of respondents replied to the question (% in agreement) |
| --- | --- | --- | --- |
| B. Specimens | Aside from whole blood, plasma and serum as first choice, what is the next ranked specimen for VBS to collect? | PMBC | 13/18 (72%) |
| D. Benefits | What is the single most important benefit to you? | Capacity building (39%) with networking and SOPs (28% each) | None reached threshold |
|  | Are there challenges to receiving benefits you selected? | Yes | 11/15 (73%) |

\*Threshold for analysis: if prioritization reached consensus ( $\geq 70\%$ of respondents rating an item $\geq 4$ and excluded if $\leq 15\%$ responded to the item or rating an item $\leq 2$ on a 1–5 scale). PBMC, peripheral mononuclear cells; SOP, standard operating procedure.

### Descriptive Analysis

#### General Interest and Biorepository Models

The VBS initiative generated high levels of interest among stakeholders, with 89% of respondents expressing either “very interested” or “extremely interested” in accessing biorepository specimens. Additionally, 85% demonstrated strong enthusiasm for involvement in biorepository operations. When asked about preferred governance models, the combined model of integrating federated standards with local governance was favored, reaching consensus in Round-1 from 78% of respondents and when repeated again at the end of the questionnaire, still maintained support of 83%, underscoring the importance of balancing standardization with the flexibility required for diverse operational contexts.

#### Specimen Prioritization

Blood specimens (whole blood, plasma, serum) were overwhelmingly prioritized for collection and sharing, with 95% of respondents ranking them as a high or very high need, The question wording related to peripheral mononuclear cells was clarified for Round-2 where it was supported by 72% of the respondents (Table 2). Longitudinal collections were emphasized, with 91% supporting serial samples from confirmed cases and 84% prioritizing pre- and post-vaccination specimens. Additionally, 82% highlighted the importance of sourcing specimens from endemic regions to ensure geographical diversity.

#### Governance and Specimen Sharing

A ranked system for specimens, based on quality and metadata, was supported by 98% of respondents. Key ranking criteria included accessibility (86%), completeness of clinical data (79%), and storage/handling conditions (76%). Barriers to specimen sharing were significant, with 76% citing international shipping challenges.

#### Sample Quality and Metadata

Standardized quality metrics were broadly supported, with 93% emphasizing the need for minimum quality standards. Metadata accompanying specimens was deemed critical. All respondents (100%) prioritized specimen descriptions (e.g., source, type), while 91% emphasized collection and handling conditions, such as temperature and freeze/thaw cycles, were highlighted by 84% reflecting the importance of robust data to support research integrity.

#### Sustainability and Services

Sustainability emerged as a pivotal concern, with unanimous support (100%) for a cost-recovery model. A sliding fee scale, adjusted based on the type of organization (e.g., academic institutions, commercial enterprises, or low- and middle-income countries), was supported by 95% of respondents. Regarding services, 79% prioritized reference panels for calibration and proficiency testing, while 77% emphasized the need for protocols and templates for specimen collection and storage. Infrastructure support, such as cold chain equipment and shipping logistics, were highlighted as essential.

#### Consent and Ethics

Ethical concerns were evident in the Delphi rounds, with only 35% affirming that broad consent as defined by the common rule in 2018[18], could be adopted within their institutions or nations for future specimen use. However, 92% supported sharing specimens for immune-based diagnostics or vaccine development. Additionally, 84% expressed interest in participating in policy development, reflecting a strong desire for transparency and collaboration in governance.

#### Institutional and Personal Benefits

Individual and institutional benefits did not reach consensus in Round-1 or Round-2. It is important to note that although 73% acknowledged challenges in specimen-sharing benefits in Round-2, no single barrier reached consensus. A lack of consensus does not necessarily indicate opposition to a particular concept. It may instead suggest that the concept is new, or poorly understood, or that respondents come from varied disciplines and cultures [18]. These variances show areas where difficult conversations may be needed with clarifications and understanding to build a VBS that effectively meets the needs of our stakeholders [19].

### Refining Priorities

In Round-2, consensus was reached on several key points:

1. **Specimen Prioritization:** Having already identified whole blood, plasma, and serum samples as high priority in Round-1 (95%), Round-2 respondents (72%) prioritized PBMC as the next type of specimen with strong support (91%) for longitudinal sampling. Additionally, 68% respondents emphasized finding solutions to ensure COVID-19 specimens are preserved for research.
2. **Barriers:** 73% acknowledged challenges in specimen-sharing benefits, with no consensus on which of the barriers carried more weight.

## Open Comments Summary

Respondents made comments on specific issues and they are list here.

### Specimen Prioritization

Respondents reinforced the prioritization of blood specimens (whole blood, plasma, serum) for collection, storage, and sharing. Other suggestions included cerebrospinal fluid, tissue, and breast milk. Feedback emphasized that specimen prioritization should adapt to specific pathogens, with strong support for longitudinal sampling.

### Barriers to Specimen Sharing

Shipping challenges were the most frequently cited obstacle (73%), compounded by infrastructure limitations, especially in regions with inadequate cold chain capacity. Regulatory complexities, such as the Nagoya Protocol [20] were also noted as a barrier to international sample sharing.

### Governance: Federated Standards with Local Adaptation

The combined governance model—integrating federated standards with local oversight— emerged as the most effective approach. Respondents highlighted the advantages of local governance in enabling rapid responses, particularly during pandemics, while federated standards ensure consistency across regions.

### Ethical and Policy Considerations

Local and national regulations posed obstacles, as identified by multiple respondents. This highlights the need for streamlined logistics and regulatory frameworks to facilitate international collaboration.

### Sustainability and Recommendations

Funding was identified as a major challenge, with respondents stressing the need for long-term coordination and infrastructure development. Recommendations included establishing training programs to build equity in lower-resourced settings and incorporating standardized operating procedures. Notably, participants suggested engaging more stakeholders from regions underrepresented in the current survey, particularly Africa.

## Discussion

The Delphi survey results emphasize the importance of a flexible yet standardized approach to biorepository governance, robust quality standards, and inclusive stakeholder collaboration. These findings will guide the next stages of developing the VBS, ensuring it is equitable, sustainable, and responsive to global research needs.

The Delphi method revealed a rich tapestry of insights about the potential of a VBS. The enthusiasm among respondents highlighted the transformative possibilities but also brought to light critical areas that must be addressed to ensure its success. One of the most striking benefits of the VBS lies in its ability to foster international collaboration. Respondents expressed optimism about how the system could break down traditional barriers in sample sharing, facilitate meaningful cross-border partnerships, and provide opportunities for training and capacity building. This is particularly exciting for under-resourced regions, where the VBS could play a pivotal role in reducing disparities and promoting equity in global health research [1].

A key strength of the system lies in its ability to connect users with biorepository expertise. By aligning operations with international standards and developing Standard Operating Procedures with quality requirements [21], the VBS can elevate the reliability of biospecimen management for infectious disease research worldwide. Respondents envisioned a future with the VBS as a cornerstone for consistent, high-quality international research.

Nevertheless, this vision comes with its challenges. A notable emergent theme was the regulatory knowledge gap across countries, highlighting the need for co-developed resources to align understanding of local and national regulations. This is particularly relevant to the application of benefits under the Nagoya Protocol, as outlined in the Convention on Biological Diversity, which specifies explicit requirements for country signatories [20]. While this lack of shared knowledge could hinder cross-border collaborations, it also offers an opportunity to develop a system that addresses these gaps through accessible guidance and coproduced documentation.

Concerns about infrastructure and resource limitations were prominent in the discussions. Stable power supplies, reliable storage solutions, and contingency plans for system failures are essential to maintaining sample integrity [21]. Sustainable funding models were another area of concern, as respondents noted the challenges of securing long-term financial support for infrastructure, supplies, and personnel. These issues underline the importance of a robust operational foundation to support the VBS’s goals.

Logistics posed another layer of complexity, particularly considering past experiences during outbreaks. Shipping delays, high costs, and cumbersome regulations were frequently cited as barriers that need to be addressed. Respondents called for innovative solutions to streamline these processes, ensuring that the movement of specimens is both timely and cost-effective. The discussions also explored the unique difficulties of operating in conflict zones or disaster-stricken areas. Strategies to improve access in such environments, such as establishing proxy centers, were proposed as potential solutions to ensure inclusivity and equity.

Ethical considerations emerged as a critical theme, with respondents advocating for careful negotiation of broad consent frameworks [22]. The balance between enabling research and respecting participants’ rights was a shared priority, with consensus around the need to limit consent to intended uses and avoid misuse by third parties [22]. Another area of concern was the power dynamics inherent in global collaborations. Smaller biorepositories often face challenges in negotiating on equal terms with larger entities, highlighting the need for governance structures that prioritize fairness and inclusion. Similarly, the importance of equitable benefit-sharing was universally acknowledged, though the variability in challenges across regions pointed to the complexity of designing solutions that work for everyone.

Finally, questions about data and sample quality underscored the importance of consistency and rigor. Ensuring accurate clinical data, maintaining high diagnostic standards, and managing samples effectively and providing timely reference materials were seen as vital to the VBS’s success [23]. These discussions point to a powerful vision for the VBS, one that could transform global health research by promoting collaboration, equity, and innovation. Innovations for safe and secure storage materials, that are free of expensive ultra-low cold storage could alleviate costs for power, equipment and shipping. Yet, the path forward requires thoughtful, inclusive strategies to address the challenges and ensure the system’s benefits are accessible to all. By fostering transparency, sustainability, and shared ownership, the VBS has the potential to become a model for how global health infrastructure can evolve to meet the needs of an interconnected world.

## Challenges and Limitations

The first Delphi round found broad support for a combined biorepository model, incorporating federated standards with local governance to optimize standardization and agility. This approach can address administrative delays inherent in federated systems while allowing for the rapid mobilization of resources necessary for public health emergencies. Consensus was reached on specimen types and quality standards, although defining the normal sample, with education about why this is critical for research and practice will be a priority for the VBS. Normal control or reference samples may bring the fewest legislative hurdles and could be an appropriate foundation for testing the intercountry sharing of specimens as they are not subject to the same restrictions as infected samples. These samples can also serve as the testing ground for defining terms and differences.

Potential limitations include selection bias from snowball sampling and challenges in achieving consensus on complex issues. Language as a barrier was discussed. We learned that other language respondents were comfortable in filling out the Delphi in English, however, respondents shared that language barriers are more challenging to overcome in oral discussions. Ongoing discussions are needed to address these logistical and ethical challenges. Benefits sharing will require investigation and collaboration locally and within the federated system.

Round-2 highlighted the need for reaching persons native to countries, rather than only expatriates assigned to the regions. Respondents expressed anticipated hesitation concerning the VBS role and structure, and these findings will guide future discussions as we continue to refine the VBS model.

## Future Directions

Guided by the Delphi findings, we have launched a three-year demonstration project involving representatives from ten founding member countries (Bangladesh, Colombia, Guatemala, Indonesia, Jordan, Malaysia, Moldova, Senegal, Sierra Leone, Tahiti, and the United States). This initiative, built around the “10×10” model—10 large-volume samples from 10 donors per site for serology testing—focuses on governance, member benefits, and a sustainable operations plan. Early efforts prioritize standardizing terminology, such as clarifying “biorepository” versus “biobank,” and defining “healthy normal” to ensure consistency across sites.

Reflecting respondents’ preferences, the project emphasizes ethical sharing, sustainable access, and locally guided sample collection, supported by a buddy system to expand membership. Contingency plans include sharing calibrated test kits and harmonized standards across sites when specimen sharing proves impractical. These coordinated efforts aim to advance the VBS as a global platform for equitable, ethical, and collaborative specimen sharing.

## Conclusions

These findings highlight the transformative potential of a VBS to address critical gaps in international biorepository efforts and the significant challenges that remain to be navigated for implementation. The enthusiasm for enhanced collaboration and capacity building aligns with the VBS’s mission to promote inclusion and equity in specimen sharing. At the same time, the complexities of infrastructure, regulation, and logistics underscore the need for a strategic and comprehensive approach. Concerns about power dynamics and data quality call for robust and respectful co-created governance frameworks and agreed quality control measures. Future efforts will focus on practical solutions, including regulatory harmonization, infrastructure development in resource-limited settings, and equitable governance structures. The 10X10 pilot work will provide actionable insights into overcoming these challenges.

## End Matter

### Dissemination

The VBS research will be shared with repository groups, policymaking agencies, regional, national, and international conferences, government agency briefing sessions, the WHO BioHub, social media sites, and on the institutional websites of co-authors and members of our advisory group. Results will be open-access and publicly available with a DOI [9]. They will be shared with all respondents ahead of publication.

### Data Sharing

All de-identified data results are contained within the body of the paper or in the appendices.

## Data Availability

Data Sharing: All de-identified data results are contained within the body of the paper or in the appendices.

https://osf.io/6kct8/

## Acknowledgments

We would like to acknowledge the organizations that have worked with us on the Reconciliation of Cohort Data in Infectious Diseases (ReCoDID), Centers for Research in Emerging Infectious Diseases (CREID), Foundation for Innovative Diagnostics (FIND), The Global Health Network TGHN, and our named Delphi respondents; Dr. John Spencer, Dr. Daniel Olson, Dr. Matthias Strobl, Dr. Katherine Loens, Dr. Kevin Messacar, Dr. Oyewale Tomori, Dr. Jane Cardosa, Dr. Ute Stöher, Dr. Jorge Munoz, Dr. Sarah Bethencourt, and Dr. Nicole Wolter.

## Author Contributions

MC, TJ, JG and AP conceptualized the document. AP and LA drafted the document. AP, LA, JG, JP, GW, ZS, TJ, and MC contributed to constructing the Delphi and co-authoring the manuscript. GW led the translation and coordination for panel members for whom English was not a first language. TJ and MC acquired funding.

## References

1 Giri J, Pezzi L, Cachay R, et al. Specimen sharing for epidemic preparedness: Building a virtual biorepository system from local governance to global partnerships. PLOS Glob Public Health. 2023;3:e0001568. doi: 10.1371/journal.pgph.0001568

2 Yadouleton A, Sander A-L, Moreira-Soto A, et al. Limited Specificity of Serologic Tests for SARS-CoV-2 Antibody Detection, Benin. Emerg Infect Dis. 2021;27:233–7. doi: 10.3201/eid2701.203281

3 Sheldon M, Grimwood R, Nahas SA. *Letter to the Editor:* Biobanks and International Initiatives Are Playing a Critical Role in Redressing Historic Inadequacies in Biosample and Data Access from Underrepresented Minority Populations. Biopreservation Biobanking. 2022;20:465–6. doi: 10.1089/bio.2022.0059

4 Pluss O, Campbell H, Pezzi L, et al. Limitations introduced by a low participation rate of SARS-CoV-2 seroprevalence data. Int J Epidemiol. 2023;52:32–43. doi: 10.1093/ije/dyac178

5 Ma X, Li Z, Whelan MG, et al. Serology Assays Used in SARS-CoV-2 Seroprevalence Surveys Worldwide: A Systematic Review and Meta-Analysis of Assay Features, Testing Algorithms, and Performance. Vaccines. 2022;10:2000. doi: 10.3390/vaccines10122000

6 Logullo P, Zuuren EJ van, Winchester CC, et al. ACcurate COnsensus Reporting Document (ACCORD) explanation and elaboration: Guidance and examples to support reporting consensus methods. PLOS Med. 2024;21:e1004390. doi: 10.1371/journal.pmed.1004390

7 Lessler J, Riley S, Read JM, et al. Evidence for Antigenic Seniority in Influenza A (H3N2) Antibody Responses in Southern China. PLoS Pathog. 2012;8:e1002802. doi: 10.1371/journal.ppat.1002802

8 Zuuren EJ van, Logullo P, Price A, et al. Existing guidance on reporting of consensus methodology: a systematic review to inform ACCORD guideline development. BMJ Open. 2022;12:e065154. doi: 10.1136/bmjopen-2022-065154

9 Price A, Abdulbaki L, Giri J, et al. Supplemental material, VBS Delphi Protocol. Available at OSF.IO/6KCT8

10 Elwyn G, Nelson E, Hager A, et al. Coproduction: when users define quality. BMJ Qual Saf. 2020;29:711–6. doi: 10.1136/bmjqs-2019-009830

11 Zuuren EJ van, Price A, Blazey P, et al. Developing reporting checklist items from systematic review findings: a roadmap and lessons to be learned from ACCORD. J Clin Epidemiol. 2024;174. doi: 10.1016/j.jclinepi.2024.111490

12 Vaughn LM, Jacquez F. Participatory Research Methods – Choice Points in the Research Process. J Particip Res Methods. 2020;1. doi: 10.35844/001c.13244

13 Likert R. A technique for the measurement of attitudes. Arch Psychol. 1932;22 140:55–55.

14 Gattrell WT, Logullo P, Zuuren EJ van, et al. ACCORD (ACcurate COnsensus Reporting Document): A reporting guideline for consensus methods in biomedicine developed via a modified Delphi. PLOS Med. 2024;21:e1004326. doi: 10.1371/journal.pmed.1004326

15 Research ethics (including CUREC). https://researchsupport.admin.ox.ac.uk/governance/ethics (accessed 29 December 2024)

16 ICMJE | Recommendations | Defining the Role of Authors and Contributors. https://www.icmje.org/recommendations/browse/roles-and-responsibilities/defining-the-role-of-authors-and-contributors.html (accessed 29 December 2024)

17 Improving the Quality of Web Surveys: The Checklist for Reporting Results of Internet E-Surveys (CHERRIES) - PMC. https://pmc.ncbi.nlm.nih.gov/articles/PMC1550605/ (accessed 29 December 2024)

18 Grubb AM, Easterbrook SM. On the Lack of Consensus over the Meaning of Openness: An Empirical Study. PLoS ONE. 2011;6:e23420. doi: 10.1371/journal.pone.0023420

19 Failure_Institute. Consensus is killing the difficult conversations. Fail. Inst. 2020. https://thefailureinstitute.com/blog/consensus-is-killing-the-difficult-conversations/ (accessed 29 December 2024)

20 Heinrich M, Scotti F, Andrade-Cetto A, et al. Access and Benefit Sharing Under the Nagoya Protocol—Quo Vadis? Six Latin American Case Studies Assessing Opportunities and Risk. Front Pharmacol. 2020;11:765. doi: 10.3389/fphar.2020.00765

21 Cornish NE, Anderson NL, Arambula DG, et al. Clinical Laboratory Biosafety Gaps: Lessons Learned from Past Outbreaks Reveal a Path to a Safer Future. Clin Microbiol Rev. 2021;34:e0012618. doi: 10.1128/CMR.00126-18

22 Maloy JW, Bass PF. Understanding Broad Consent. Ochsner J. 2020;20:81–6. doi: 10.31486/toj.19.0088

23 Haselbeck AH, Im J, Prifti K, et al. Serology as a Tool to Assess Infectious Disease Landscapes and Guide Public Health Policy. Pathogens. 2022;11:732. doi: 10.3390/pathogens11070732

